# Estimating RSV seasonality from pandemic disruptions: a modelling study

**DOI:** 10.1101/2022.06.18.22276591

**Authors:** Fabienne Krauer, Tor Erlend Fjelde, Mihaly Koltai, David Hodgson, Marina Treskova-Schwarzbach, Christine Harvey, Mark Jit, Ole Wichmann, Thomas Harder, Stefan Flasche

## Abstract

**Background:** Respiratory syncytial virus (RSV) is a leading cause of respiratory tract infections and bronchiolitis in young children. The seasonal pattern of RSV is shaped by short-lived immunity, seasonally varying contact rates and pathogen viability. The magnitude of each of these parameters is not fully clear. The disruption of the regular seasonality of RSV during the COVID pandemic in 2020 due to control measures, and the ensuing delayed surge in RSV cases provides an opportunity to disentangle these factors and to understand the implication for vaccination strategies. A better understanding of the drivers of RSV seasonality is key for developing future vaccination strategies.

**Methods:** We developed a mathematical model of RSV transmission, which simulates the sequential re-infection (SEIRRS4) and uses a flexible Von Mises function to model the seasonal forcing. Using MCMC we fit the model to laboratory confirmed RSV data from 2010-2022 from NSW while accounting for the reduced contact rates during the pandemic with Google mobility data. We estimated the baseline transmission rate, its amplitude and shape during RSV season as well as the duration of immunity. The resulting parameter estimates were compared to a fit to pre-pandemic data only, and to a fit with a cosine forcing function. We then simulated the expected shifts in peak timing and amplitude under two vaccination strategies: continuous and seasonal vaccination.

**Results:** We estimate that RSV dynamics in NSW can be best explained by a high effective baseline transmission rate (2.94/d, 95% CrI 2.72-3.19) and a narrow peak with a maximum 13% increase compared to the baseline transmission rate. We also estimate the duration of post infection temporary but sterilizing immunity to be 412 days (95% CrI 391-434). A cosine forcing resulted in a similar fit and posterior estimates. Excluding the data from the pandemic period in the fit increased parameter correlation and yielded less informative posterior distributions. The continuous vaccination strategy led to more extreme seasonal incidence with a delay in the peak timing and a higher amplitude whereas seasonal vaccination flattened the incidence curves.

**Conclusion:** Quantifying the parameters that govern RSV seasonality is key in determining potential indirect effects from immunization strategies as those are being rolled out in the next few years.

## Introduction

Respiratory syncytial virus (RSV) is an endemic virus and a leading cause of acute lower respiratory tract infections in young children. The majority are infected at least once before they are two years old [1]. Subsequent re-infections and infections at older age are thought to result in less severe disease [2,3]. Typical complications of RSV in young children are bronchiolitis or pneumonia. The proportion asymptomatic among infected individuals ranges from 9% in <1 year olds to 78% in adults [4].

To date, the only available measure for pharmaceutical prevention of severe disease is palivizumab, a short-lived and costly monoclonal antibody; no active immunisation product has been licensed yet. Several vaccine candidates and a long-lasting monoclonal antibody are undergoing trials in young infants, pregnant women or the elderly [5,6]. The optimal vaccination strategy depends on the efficacy and duration of immunity conferred [7]. Anticipating short-lived protection from immunisation, seasonal immunisation of infants before the onset of RSV season was estimated to be more cost-effective than year-round vaccination [6,8].

RSV has a distinct and consistent seasonal pattern in many temperate climate settings [9,10]. The regular seasonality is likely an interplay of the seasonal fluctuations in human contacts, meteorological determinants that govern pathogen viability and host susceptibility, as well as the characteristics of the immune response following infection [11–16]. The factors that govern seasonality have been challenging to quantify, and a given seasonal disease pattern may be modelled with various combinations of parameter values. Identifiable and robust parameters are crucial for predicting incidence and determining the potential indirect effects from immunisation strategies as those are being rolled out in the next few years.

The COVID pandemic, where mobility was restricted globally and contacts were reduced dramatically while seasonal climatic pathogen properties and duration of RSV immunity were unchanged, provides a unique opportunity to estimate the magnitude of the different factors contributing to the regular RSV seasonal pattern observed in temperate settings. The COVID control measures have disrupted the seasonal pattern of RSV around the globe. Countries in the Southern hemisphere, where the initial lockdowns occurred at the beginning of the season, showed an abrupt termination of the 2020 season followed by a delayed peak once the restrictions were eased (see e.g. Australia [17–19] or South Africa [20]). In the Northern hemisphere, the 2020/2021 season following the lockdown period also exhibited a peak delayed by several months in multiple countries (see e.g. France [21], Israel [22], USA [23], Japan [24], UK [25] or Spain [26].

In this study, we use incident RSV data from New South Wales (NSW, Australia) from sentinel laboratories from 2010-2022 and fit a dynamic transmission model to quantify the magnitude of RSV seasonality and the duration of immunity taking into account the reduction of contacts during the pandemic. We hypothesised that the additional information in the data resulting from the contact reductions facilitates parameter inference by constraining the parameter space. To approximate the contact reduction, we use Google mobility data. We then compare the resulting parameter estimates to a fit to a subset of pre-pandemic data and to a fit using an alternative transmission forcing function. Finally, we investigate how two hypothetical vaccination strategies may impact the regular seasonal pattern.

## Methods

### Data

We used two types of RSV data to inform our model parameters. The main data source was an incidence times series from the respiratory diseases surveillance system in New South Wales [27,28]. These reports comprise the weekly incidence of laboratory confirmed RSV and influenza cases, as well as the total number of tested cases (influenza only). The data include only symptomatic patients that sought healthcare and had a sample taken, which are an unknown fraction of the total number of cases. RSV cases are defined as a patient for whom RSV was detected by cell culture, nucleic acid testing (PCR), antigen testing or serology (https://www1.health.gov.au/internet/main/publishing.nsf/Content/cda-surveil-nndss-casedefs-cd_rsv.htm). We included cases recorded between 8. January 2010 and 6. February 2022.

From 2015 - 2019 both the base levels and the amplitude of RSV notifications increased, which coincided with an increasing number of samples tested for influenza in the same surveillance network (see Figure S1a). However, emergency department visits for bronchiolitis remained roughly the same between 2010-2019 with around 350 cases per week at peak seasonal activity (Figure S1b) suggesting an increase in sensitivity of the sentinel surveillance rather than an increasing burden of disease. The increase in testing was in part due to the introduction of new multiplex PCR assays in some hospitals in 2015 [29]. Secondly, we used annual RSV infection attack rates estimates from a longitudinal, prospective study in Kenyan households, which collected biweekly samples of all participants regardless of symptoms [4]. The change in contact patterns due to COVID-19 mitigation measures since early 2020 was approximated through the Google Community Mobility index [30]. We selected the mobility index at workplaces as most indicative of changes in RSV transmission relevant contacts besides schools (which are not covered in the Google mobility data). We used the rolling 7-day median index to smooth the data while retaining changing trends in mobility (Fig. S2a).

### Model and parameter calibration

We developed a dynamic compartmental transmission model with sequential acquisition of immunity (adapted from [6,14]) to account for the natural history of RSV infection. Briefly, individuals are classed into five different states (S, E, I, R^1^, R^2^) at four different levels (0,1,2,3+) that reflect the number of previous RSV infections. The two R compartments at each level correspond to an Erlang distributed duration of immunity [31]. Infants are born into the susceptible compartment *S*_0_ at a rate *μ*. When infected, they become latently infected (*E*_0_) and subsequently infectious (*I*_0_) after which they become temporarily but fully immune (*R*_0_^1^ followed by *R*_0_^2^). After this temporary but sterilising immunity wanes, individuals become partially susceptible again (*S*_1_). Subsequent re-infection will lead to similar progression while keeping track of the number of previous infections until an individual has had three infections, after which infections are assumed to no longer lead to differences in immunity and susceptibility. Susceptibility to re-infection and duration of infectiousness are assumed to decrease with repeat exposure. The duration of the protective immunity was assumed to be the same at each level of immunity. The population was modelled with a life expectancy of 80 years and a constant birth/death rate *μ*, which results in a closed population of constant size. We assumed that the effective per capita transmission rate (*β*_*eff*_) varied seasonally due to seasonal differences in contacts and pathogen viability in the environment. We modelled the seasonal forcing with a modified Von Mises function (MVM) with parameter *k* that allows adjusting the variance of the peak (see supplement). The MVM can take any shape between a cosine-like function and a flat line with a sharp peak on a single day. The change in contact behaviour during the pandemic was modelled by scaling the force of infection with a time-varying parameter *i* with *i* ∈ [0,1], which was estimated from Google Community Mobility data [30] (see supplement).

We fitted the model jointly to the two datasets described above. We assumed that the reported weekly incidence is a fraction *ρ* of the incident symptomatic cases, and that only cases of levels 0-2 would lead to severe enough disease requiring testing. The present surveillance data does not provide age information but earlier laboratory data confirm that >93% of all tests were for children aged <5 years old [32,33]. The reporting rate comprises the overall combined probability of symptomatic disease, visiting a GP and having a sample taken and analysed by the laboratory. To account for the non-linear increase in sampling and testing over time, we assumed *ρ* followed a sigmoid function with two parameters: an initial reporting rate *ρ* _0_, and a rate of increase, *q* (see supplement). For (2), we assumed an average level-specific attack rate calculated as the cumulative cases at the end of the year divided by the population size at the beginning of the year for a given level of reinfection. The model was fitted jointly to the reported weekly incidence of cases assuming a Negative Binomial likelihood with dispersion parameter *ψ*, and to the yearly attack rate data from a longitudinal Kenyan household study assuming a Binomial likelihood (see supplement).

To improve the identifiability of all parameters and convergence, we fixed all parameters for which we had acceptable point estimates and which did not depend on a specific geographical setting to values from published literature. The following parameters of interest to the study question were estimated: the baseline transmission rate (*β*_0_), the amplitude (*η*), peak width (*k*) and phase shift (*φ*) of seasonal transmission rate and the reduction of the susceptibility to reinfection at the different levels (*δ*_1_, *δ*_2_, *δ*_3_) (Table 1). Moreover, we estimated the baseline and rate of increase in the reporting rate (*ρ* _0_ and *q*) and the dispersion parameter of the observation model (*ψ*), since these depend on the data and cannot easily be compared between settings. The proportion asymptomatic or the duration of infectiousness are often assumed to differ by age and/or level of reinfection, but the latter is not commonly measured in observational studies. We therefore converted age-specific estimates to level-specific estimates assuming that level 0 corresponds to age <1 year, level 1 to age 1-2 years, level 2 to age 3-5 years and level 3 to 5+ years.

**Table 1.**
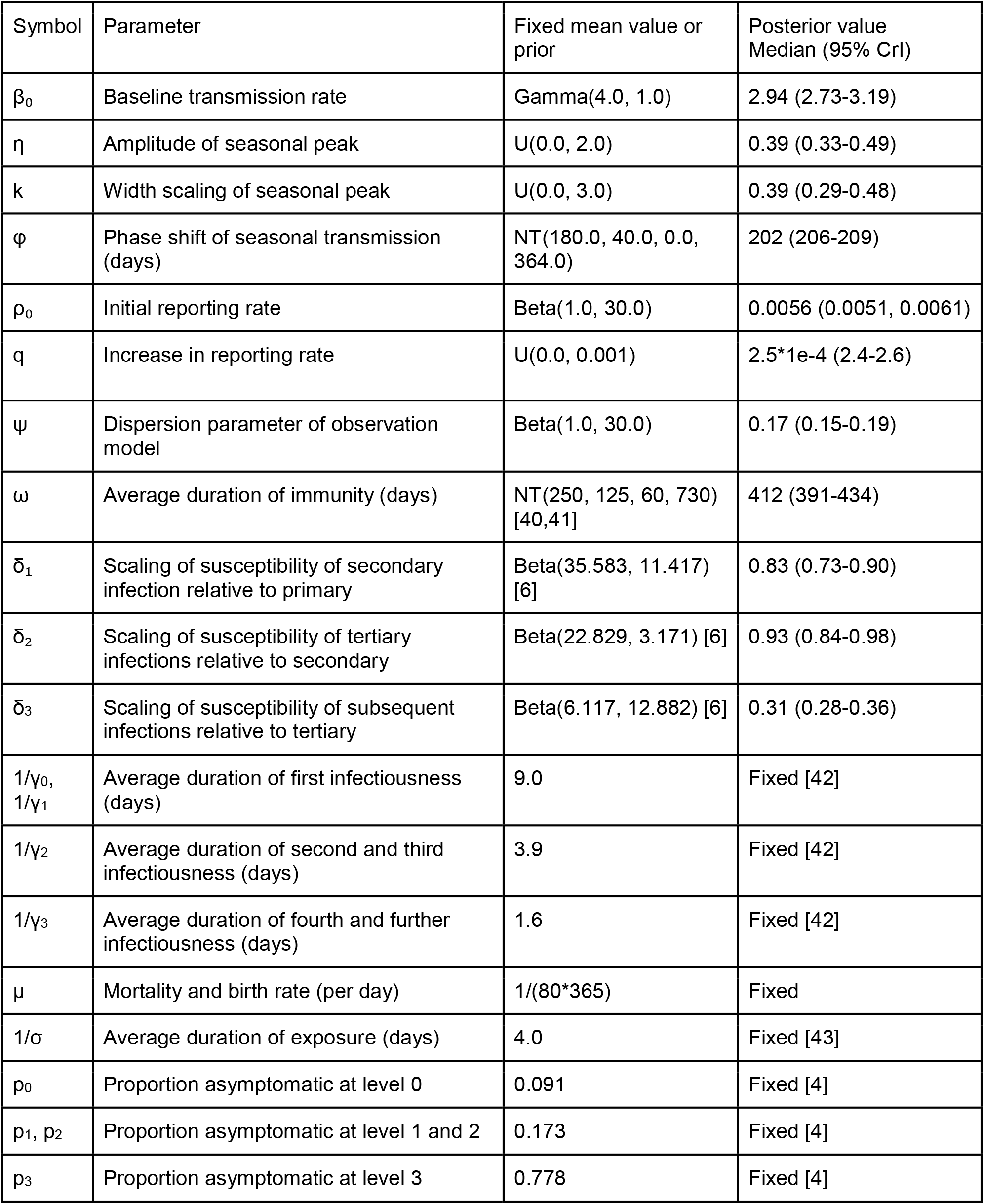
Parameters used in the model. Values were either fixed from literature or fitted. U=Uniform(lower bound, upper bound), NT=normal truncated(mean, sd, lower bound, upper bound).

The model was fitted with a Bayesian MCMC algorithm with the priors shown in Table 1. We simulated the model with a run-in period of 100 years before evaluating the likelihood to ensure a stable periodic orbit (quasi equilibrium). We used the Hamiltonian Monte Carlo (HMC) No-U-Turn Sampler (NUTS) algorithm implemented in the Julia package Turing.jl [34,35] to run 10 chains with 500 iterations burn-in and 500 posterior samples per chain. Convergence was confirmed with the Gelman-Rubin statistics (estimates <1.1 were considered indicative of convergence). The uncertainty of model-derived quantities was calculated from the 2.5th and 97.5th percentile of all trajectories over each modelled data point (posterior predictive interval PPI). Marginal posteriors are reported as median and 2.5th - 97.5th percentiles Credible Intervals (CrI).

### Sensitivity analysis

To explore the sensitivity of the model parameters to changes in the underlying data, we fit the same model to a subset of the RSV incidence data covering only the pre-pandemic period 2010-2019, and compared the posterior cross-correlations and the marginal posterior distributions. We also studied the effect of the choice of the seasonal forcing function on the trajectory fit and the posterior parameter estimates in a sensitivity analysis. We re-fit the model to the 2010-2022 data using a cosine forcing (see supplement), which has one parameter less and produces a symmetric peak and nadir around an average transmission rate. The trajectory fit was compared to the main results with the Von Mises forcing using Pareto-smoothed importance sampling leave-one-out cross-validation (LOO) [36]. Finally, we omitted the Kenyan attack rate data from the likelihood function and fitted the model with the Von Mises forcing only to the time series data from NSW, and compared the results.

### Vaccination strategies

To investigate how active immunisation of infants against RSV may interfere with RSV seasonality, we used the posterior estimates to forward simulate the pattern of RSV seasonality in NSW and its changes under two different vaccination strategies. We simulated both continuous (year-round) and seasonal vaccination in previously unexposed, fully susceptible individuals in level 0 as a proxy for vaccinating infants. The model equations are given in the supplement. Seasonal vaccination was simulated with a time window of seven months during RSV season (March-October), corresponding to the seven months of RSV season when monoclonal antibodies are commonly administered in NSW [37]. For simplicity we assume that eligible individuals who are in the exposed (*E*_0_) or infected (*I*_0_) state before vaccination would not generate an additional immune response following vaccination but simply progress to the next regular level of immunity. We also assume that the vaccine confers the same type and duration of immunity as the first infection. A temporary, sterilising immunity has been observed in trials for a live-attenuated vaccine in children [38] and a protein-based vaccine in adults [39] From the simulated trajectories we summarised 1) the shift in peak activity, 2) the changes in the amplitude and 3) the yearly percentage of cases averted compared to the baseline scenario of no vaccination. The simulated effective vaccination coverage ranged from 10% to 100% in 10% steps. The effective coverage is to be interpreted as the product of distribution coverage and vaccine effectiveness. We did not include the passive immunisation with monoclonal antibodies in our model, which prevents severe disease but does not lead to sequential acquisition of immunity. The number of infants receiving the currently licensed monoclonal antibodies is small with respect to the total population of infants and we deem the effect of a delayed susceptibility in these infants irrelevant for the transmission dynamics in our model.

## Results

### RSV seasonality

In the 10 years prior to the COVID-19 pandemic, RSV in NSW commonly peaked mid-July (median ISO week 28, range ISO week 18-32, i.e. May to August) with 66-79% of all annual cases occurring during the five months between April and August (Fig. S1C). In 2020, the beginning RSV season was abruptly terminated in March followed by a delayed peak in December 2020 (6 months after the typical season) and a smaller regular peak in July 2021 (Fig. S1D).

The model was able to replicate the regular seasonal pattern, the increase in reported incidence over time as well as the pandemic-related disruption with the exception of the first half of 2021, where the model overestimated the reported cases (Fig. 1A & B). We estimated RSV transmission to be high but only moderately seasonal with 2.94 (95% CrI 2.72-3.19) effective contacts per day for most of the year and a narrow peak with a maximum of 3.33 (95% CrI 3.04-3.69) effective contacts per day (13.1% increase above baseline, 95% CrI 12.7-13.6%) at week 29 at the end of July (Figure 1C). The average duration of sterilising, post exposure immunity, the other determinant of regular seasonal RSV behaviour, was estimated as 412 days (95% CrI 391-434 days) (Table 1). We also estimated that 0.6% (95% CrI 0.5-0.6%) of symptomatic RSV infections among levels 0-2 were reported through the sentinel surveillance in 2010, increasing to 15.3% (95% CrI 14.5-16.2%) in 2022 (Fig. 1D). The level-specific attack rates were 63% (95% PPI 60-66%), 59% (95% PPI 57-62%), 58% (95% PPI 56-61%) and 35% (95% PPI 33-36%) for levels 0-3, respectively, which closely matches the attack rate data the model was fitted to (Fig. 1E). Overall, we estimate that the average incidence was ∼2.9 Million cases (95% PPI 2.8-3 million) per year (36% of the population). MCMC trace plots, diagnostics and additional numerical results are shown in the supplementary text and Figures S3A, S4A.

**Fig. 1.**
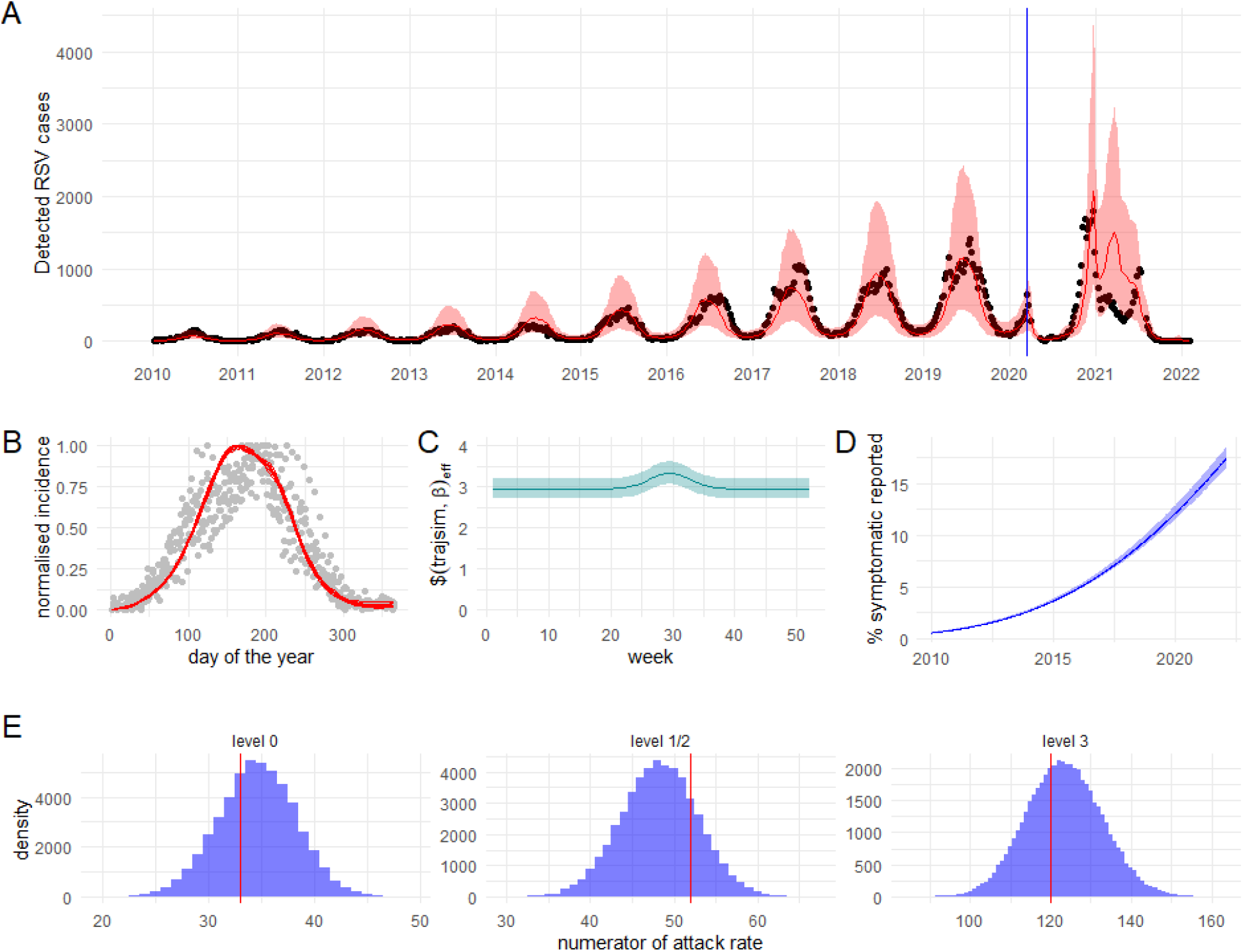
Fitted model. The shaded areas represent the 95% posterior predictive intervals; the darker lines represent the medians. A) Simulated trajectory fit and the weekly data points (black dots). The blue vertical line denotes the first implementation of the lockdown measures in NSW. B) The normalised fitted total incidence (red line) matches the normalised observed data (grey dots) except for the peak timing, which is likely preceded by a few days. C) The fitted seasonal transmission rate (*β*_*eff*_) is large with a narrow peak in July and a maximum increase of 1.13 times the baseline. D) The fit also suggests an initial reporting rate of <1% of all symptomatic cases in children increasing to 15% in 2022. E) The numerator of the attack rates as fitted by the model (blue distribution) matched the data points (red line).

### Sensitivity analysis

Fitting the same model to the pre-pandemic period (2010-2019) resulted in seemingly bimodal posterior samples (Fig. S3B) due to two chains getting stuck in a local mode as suggested by the log posterior plot (Fig S4B). For the estimation of the posterior predictive and the parameter correlation we therefore excluded these two chains. The fit to the time series data was almost identical to the result from the full dataset (Fig. S5A). However, several of the parameters that govern seasonality, particularly the baseline effective transmission rate (*β*_0_) and the amplitude of the seasonal forcing (*η*) were highly positively correlated (Fig. S5B top) suggesting a poor parameter identifiability. Including the pandemic period in the fit successfully reduced the correlation among most of the parameters with the exception of a negative correlation between *η* and *k* (which defines the width of the seasonal forcing) (Fig. S5B bottom). It’s likely that the model fits best with a specific area under the curve of the transmission rate during the season. Fitting to the full data set also resulted in narrower and more informative posteriors (Fig S5C).

Our main findings of a low seasonal forcing for RSV were robust to the shape of the forcing function. When the seasonal forcing was modelled with a cosine function, the model fitted the data almost as well (expected log pointwise predictive density -3485.94) as with a Von Mises function (ELPD -3413.37) but produced a larger uncertainty (S6A). The effective transmission rate was slightly lower at the nadir (2.82, 95% CrI 2.64-3.02), and the amplitude (maximum over minimum) was 1.10, which is a similarly weak seasonal forcing as with the Von Mises function (1.13) (Fig. S6B). The posterior densities were similar for the two forcing functions (Fig. S6C). However, the duration of immunity was estimated to be slightly shorter for the cosine model (392 days, 95% CrI 371-414 days). Of note, the cosine model turned out to be more challenging to fit. The majority of the chains did not converge within 48 hours and had to be cancelled, which could mean the posterior samples we obtained (Fig S3C, Fig. S4C) may not be representative of the target distribution.

Fitting only to the time series data from NSW, omitting the Kenyan attack rate data, resulted in a poorer chain mixing (including one chain stuck in a local mode) (Fig. S3D) and longer runtime, which, when using HMC NUTS, is indicative of a more difficult posterior to sample from. The trajectory fit was almost identical to the fit from the joint datasets (Fig. S7A), and the attack rates predicted by the model for the levels 0-2 were similar (Fig S7B). However, the predicted attack rates at level 3 were marginally higher when fitted without the AR data (median 41%, 95 % CrI 36-46%). Including the additional data for the attack rate led to more identifiable parameters (Fig S7C) and narrower marginal posterior distributions for some parameters including the duration of immunity (*ω*) (Fig. S7D). These results suggest that including the additional attack rate data improved our inference framework.

### Impact of vaccination strategies on seasonal pattern

We found that despite the relatively weak seasonal forcing, neither continuous (year-round) or seasonal immunisation of infants is likely to disrupt RSV seasonality as substantially as the COVID-19 pandemic did. Continuous vaccination is predicted to lead to delayed and more pronounced seasonal patterns with a narrower peak at all levels of (re-)infection (Fig. 2) with a higher peak at lower coverages. Seasonal vaccination on the other hand flattened the seasonal peak but increased the incidence off-season. Seasonal vaccination at 100% coverage led to a weak bimodal pattern in those previously infected with a peak at the beginning and the end of the regular season. Due to the more condensed season caused by continuous vaccination, the infection incidence at peak can increase by up to 156% in the previously unexposed and up to 200% in previously exposed individuals if vaccine coverage is low (Fig. 3A). Only if vaccine coverage exceeds 50% the peak incidence among previously uninfected decreases compared to no vaccination. The peak timing is always delayed under a continuous strategy (Fig. 3B). The delay increases with increasing vaccination coverage with a maximum delay of 80 days in the previously exposed at 100% coverage. Seasonal vaccination also delays the peak at level 3 by up to 17 days at low coverage, but generally advances the peak at all levels up to 129 days at 100% coverage. Both continuous and seasonal vaccination prevented a similar amount of cases (Fig. 4). In the previously unexposed (level 0) the proportion of cases prevented increased roughly linearly with increasing coverage and with only minimal displacement of infection and disease into other groups (maximum 0.2%).

**Fig. 2.**
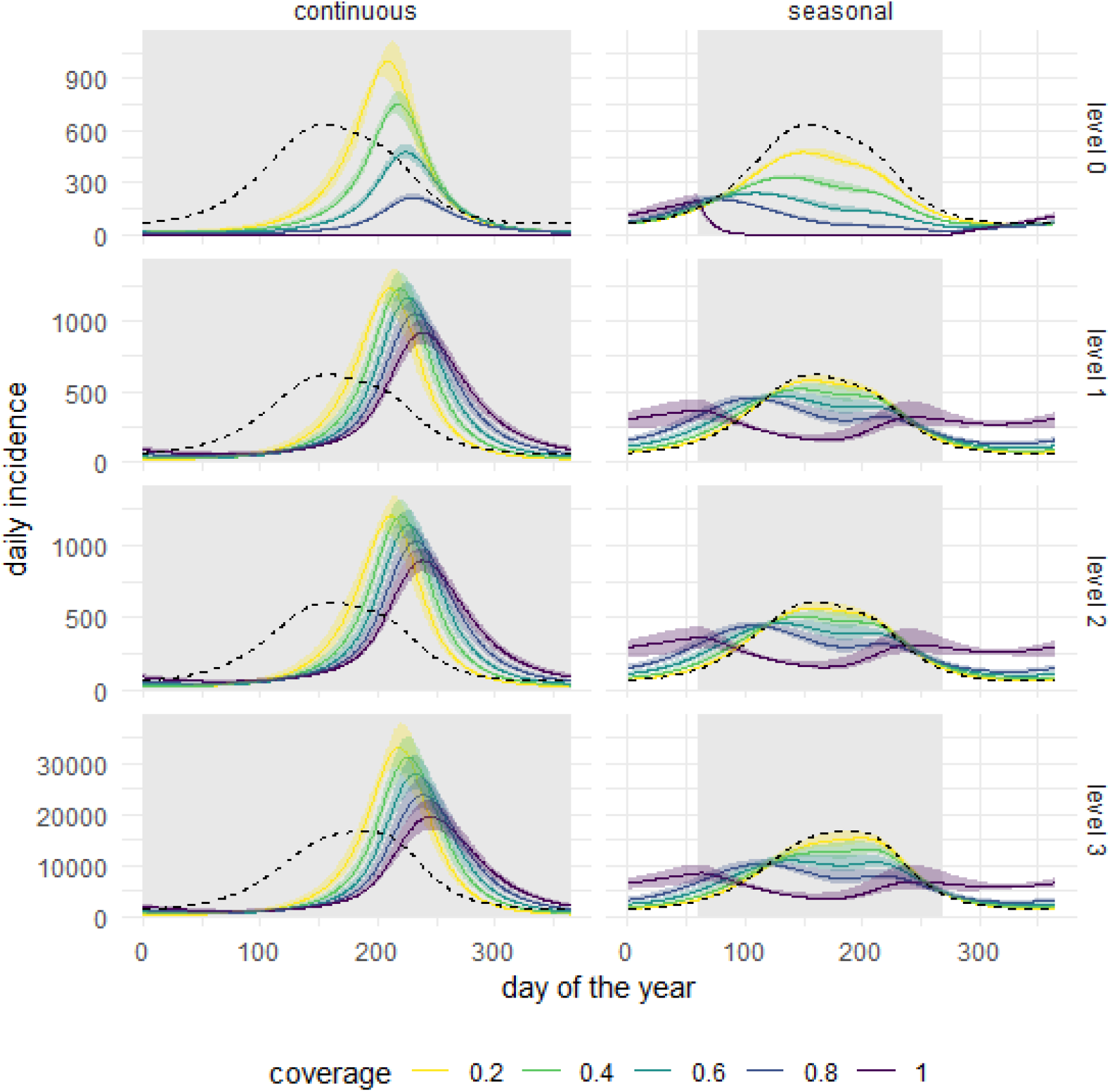
Temporal changes in seasonal RSV incidence under different vaccination coverages and strategies (seasonal vs. continuous, vaccination windows indicated by grey shaded areas) compared to no vaccination (black dashed lines). Continuous vaccination leads to more narrow epidemic peaks with larger amplitude and delayed peak timing at all levels for all coverages (except 100% coverage at level 0). Seasonal vaccination at coverages less than 100% flattens the epidemic peak at all levels. 100% seasonal coverage leads to two small peaks at the beginning and the end of the regular season for levels 1-3.

**Fig. 3.**
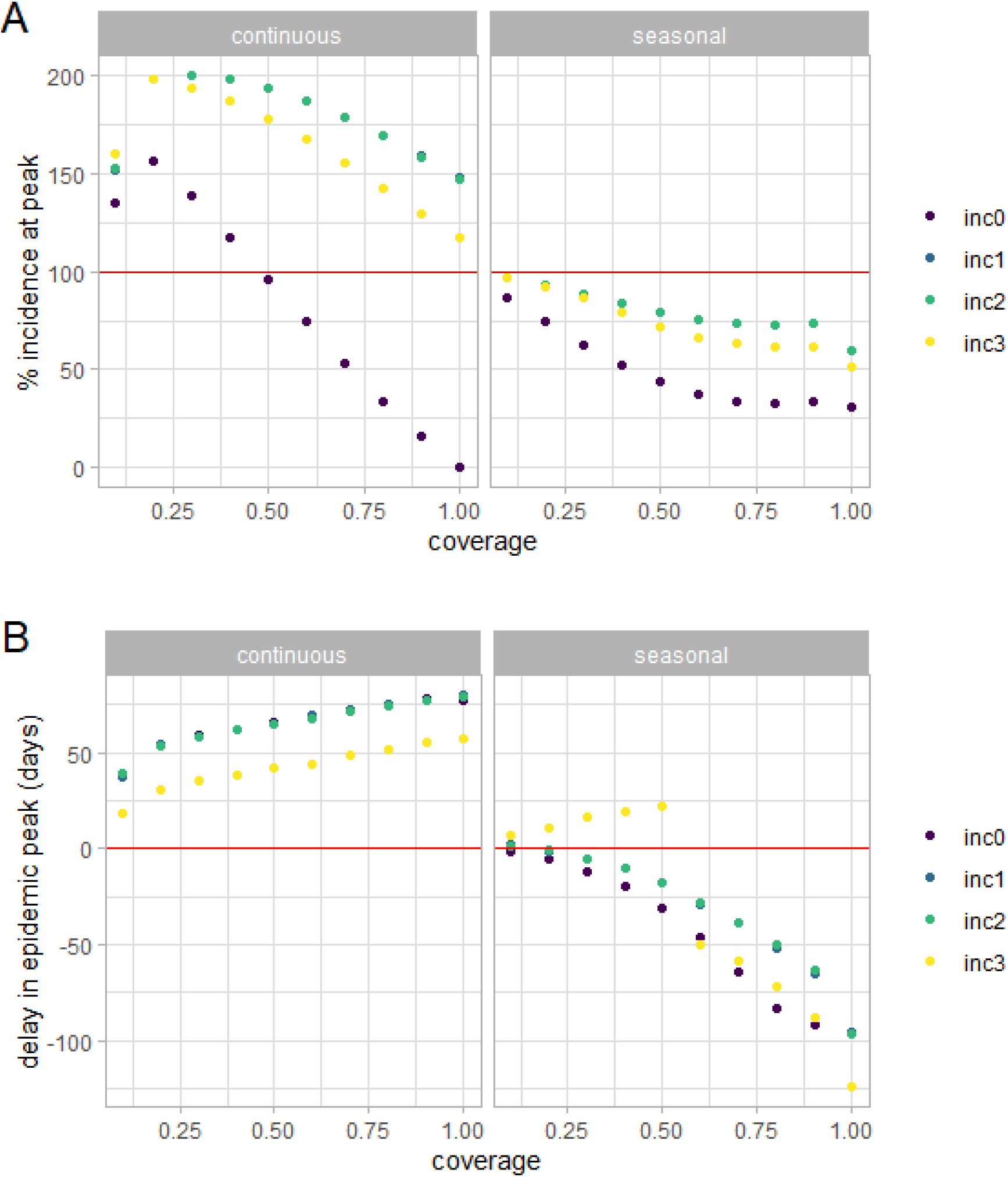
Quantitative effect of different vaccination coverages on peak timing and amplitude at all levels (0-3). A) Percentage change of incident cases at peak relative to no vaccination. Continuous vaccination increases the peak amplitude for level 0 if coverage is below 50%, and for levels 1-3 regardless of coverage. Seasonal vaccination always decreases peak amplitude B) Continuous vaccination delays the peak for all levels and coverages with a maximum of 80 days at coverage of 100%. Seasonal vaccination delays the peak for level 3 with a maximum of 17 days at a low coverage but otherwise advances the peak for all levels with a maximum of 129 days at 100% coverage.

**Fig. 4.**
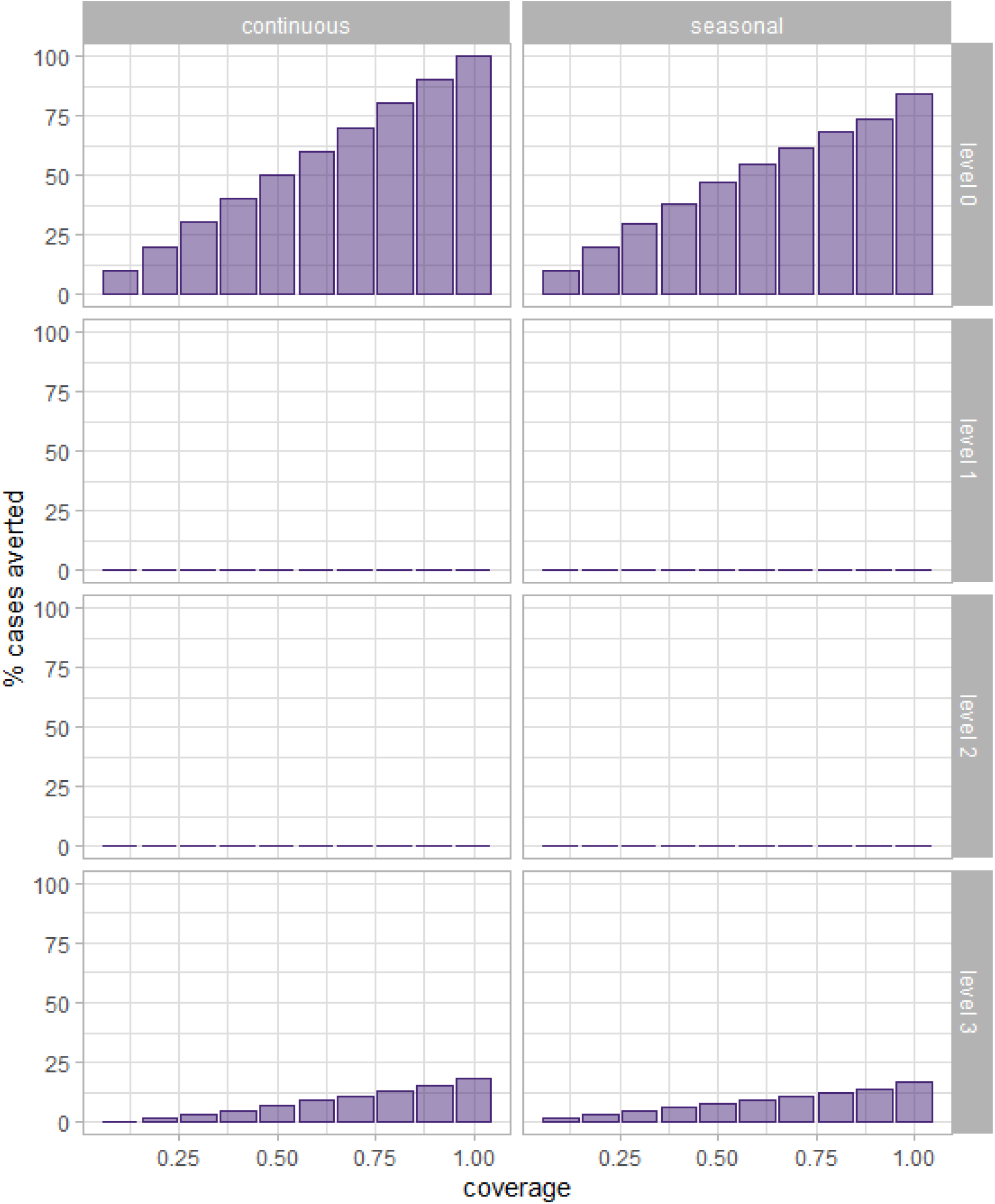
Percentage of cases averted by strategy for each level of (re-)infection. The prevention of cases at levels 0 and 3 increases linearly with coverage, and continuous and seasonal vaccination prevent almost the same percentage for a given coverage. Vaccination shifts more fully susceptible individuals to level 2 than infection alone leading to a small increase in cases at levels 1 and 2 at high coverage (maximum 0.2%, not visible on the graph).

## Discussion

In this study, we show how pandemic disruptions of RSV can be used to quantify the factors that drive the seasonal pattern. In a setting like NSW, the best fitting transmission rate is high but mostly constant throughout the year with a single, narrow peak in winter with a maximum amplitude of 1.13 times the minimum. Our sensitivity analysis confirmed our suspicion that fitting only to the pre-pandemic time series resulted in an acceptable match to the time series data but also in a correlation of the parameters governing seasonality. Excluding the attack rate data also led to higher overall parameter correlation and less informative posteriors. When the transmission rate was modelled as a cosine function, we could also obtain a similarly good fit to the data as with the Von Mises function (albeit with a wider uncertainty) and similar posteriors. However, the lower transmission rate was counteracted by a lower duration of immunity. The low amplitude of seasonal forcing (maximum over minimum) estimated in both models (cosine and Von Mises) suggests that moderate changes in contact rates and/or pathogen viability are sufficient to trigger a seasonal peak. The amount of seasonal forcing estimated in other studies is quite heterogeneous (ranging from 1.04 to 9.8 times the baseline [6,40,44–54], which may suggest problems in the parameter inference but also geographical heterogeneity in the seasonal transmission parameter. This also emphasises the need for more data resulting from quantifiable disturbances of the regular RSV dynamics from other geographical settings to narrow down the parameter space. Not only the seasonal forcing but also the average duration of infection induced immunity in the literature - either as a fitted model parameter or estimated from observational studies - varies substantially (range 169 - 744 days [6,40,44–46,48,49,51–53]. Here we show that based on our model, the immunity may last longer than the ∼250 days suggested by Australian reinfection data [40]. The high force of infection and the resulting low average age at first and second infection is also consistent with data from a longitudinal study from Finland, which shows that ∼60% of children under the age of three years were re-infected every year [55].

Our simulation study shows that a strategy of continuous vaccination of previously unexposed individuals can lead to a delayed and more extreme seasonal pattern (both among the previously unexposed and the total population) particularly for low coverages. Continuous vaccination reduces the pool of susceptibles (level 0) leading to a low level of RSV circulation. When the total availability of susceptibles is large enough, a seasonal outbreak with a narrow and large peak is possible. These potential changes in the amplitude and peak timing of RSV following vaccination have practical implications for health services and public health strategies. Even though year-round vaccination averts cases, it may lead to undesirable disease dynamics. A large seasonal surge in paediatric cases as a result of low but continuous coverage could lead to an excess of paediatric ICU admissions or attendances to other health services, which in the worst case could result in higher paediatric mortality. Seasonal vaccination on the other hand flattens the seasonal peak in our model at most coverages, which leads to more manageable disease dynamics. However, the increase in off-season incidence would require more year-round vigilance and adjusted testing guidelines in both infants and elderly. On a more theoretical level, our results also underline the need to investigate the mechanistic drivers of seasonality more in depth. In our model, the seasonal forcing of the transmission rate (*β*_*eff*_) can be interpreted as a function of the seasonally varying contact rate of people and the seasonally varying probability of transmission upon contact. Our model fit suggests a narrow peak of the seasonal forcing fits slightly better than a symmetric cosine-like behaviour for a setting like NSW. Indoor contact rates are crucial for diseases transmitted via aerosols, and the time spent indoors has a clear seasonal variation. The seasonality of indoor activities depends on the climate zone. As shown by a recent study, seasonal indoor activity in the Southern USA (which includes the same Köppen Geiger climate zone as NSW) showed a rather narrow peak in Winter and did not fit well to a cosine function [56]. Further research to disentangle and quantify the magnitude of seasonal contacts, partial host susceptibility, pathogen viability and the association with seasonally-determined factors such as meteorological conditions, school terms, time spent indoors and periodic social gatherings could provide further insight into the seasonality of RSV, and lead to more identifiable and realistic model parameters tailored to specific settings.

Our study has limitations and uncertainties. Firstly, our model is not age-structured, which required us to make some simplifying assumptions about immunity and vaccination. In the continuous vaccination strategy, we assume that the never exposed, vaccinated group are babies who are vaccinated at birth. In the seasonal strategy, only those never exposed and those currently infected are vaccinated. In practice, children who are immune after infection for the first time cannot readily be identified as being immune without serology testing, and will therefore likely also be vaccinated, which may or may not further improve their immunity and decrease their susceptibility, which could influence the transmission dynamics at a population level. The active immunisation of babies at birth is likely not feasible either, and year-round vaccination will probably be done during routine check-ups in the first year of life. Our model also does not consider maternal antibodies in new-borns, which provide some level of protection during the first months of life. This type of passive immunisation depends on the number of women that are pregnant in the last trimester and recovering from RSV. A more structured model that includes age and differential contact patterns may be needed to adequately track these numbers throughout the year. An age-structured model may also improve the prediction of the incidence following vaccination because age-specific contact patterns give a better approximation of the FOI than the infection-level specific FOI in our current model. Other factors such as seasonal birth pulses [57] are known to influence the periodicity of childhood infectious diseases, but the birth rate of NSW is remarkably constant throughout the year, and thus has little to no impact on the seasonal RSV pattern in NSW. We also did not include the seasonal migration of workers since we did not have access to migration data and do not know enough about the mixing pattern with the general population.

Secondly, the data we use for parameter calibration have their limitations. The attack rate data are taken from a prospective household study in Kenya where circulation of RSV may be different from NSW. A prospective study in Australia showed an attack rate of 35% in 0-1 year olds and 60% in 1-2 year olds [58]. The second estimate agrees well with the Kenyan data, while 0-1 year olds seem more protected in Australia, possibly due to maternal antibodies, which we have not modelled here. No data were available from Australia for older children or adults. Limited data from various high income settings suggest that the symptomatic attack rates in adults may be in the range of 3-11% [59–61], which - assuming that ∼78% of adults are asymptomatic [4] - corresponds roughly to an attack rate of 13-50%. A 20% overall attack rate was observed among women participating in the placebo group of a clinical trial [62]. A better understanding of the true level of circulation of RSV by age group and geographical setting would improve not only our parameter estimation but also any predictions for vaccination strategies. We also made some assumptions about the laboratory data, which represent only a small and unknown fraction of all cases. The sigmoidal increase in detection and reporting is an approximation we chose in the absence of the number of RSV tests performed. While the sigmoid function seems to describe the pre-pandemic increase well, the reported RSV incidence during the first half of 2021 was not matched very well by the model. It is conceivable that RSV testing was reduced during the pandemic compared to 2019, but we could not quantify the true time-varying underreporting with the type of data available to us. Additional seasonal variation in reporting, e.g. increased detection of cases with disease exacerbation due to seasonal pollution [63] or increased testing during RSV season because of a higher expectation, cannot be ruled out. Additional data such as bronchiolitis emergency department visits, which are routinely collected and reported and do not depend on changes in testing platforms, could help to further narrow down the parameter estimates of our model.

Thirdly, Google Community Mobility data may not be the optimal proxy for the overall reduction in contacts during the COVID-19 pandemic. A limited number of surveys conducted in Australia between April and July 2020 showed that the average number of daily non-household contacts in NSW were reduced by 70% during the initial phase and remained reduced by 25% in July [64]. The fitted contact reduction based on these survey data correlates with the mobility reduction, but the Google data may underestimate the actual reduction by up to 20% (Figure S2b). To our knowledge, there are no data on the magnitude of contact reduction after July 2020 for Australia. In Germany, Google mobility data were found to correlate well with the contact data collected in 2020 when weighted for home/non-home contacts [65]. A similar 1:1 correlation of the reduction of mobility at workplaces with the reduction of work contacts was also found in the UK with the CoMix study (see Fig. S1 of [66]). Given the good correlation found in these other studies and the lack of individual contact data for 2021, we considered the Google Community Mobility data an acceptable proxy measure for modelling reduced transmission. A better understanding of how contact rates related to mobility during the pandemic in Australia would be helpful in narrowing down the posteriors and further improving the fit during the pandemic phase. It is possible that the increase in mobility in 2021 compared to the previous year is not a good indicator of an increase in effective contacts because of continued measures such as mask wearing, and the mismatch between the model and the data during the first half of 2021 is the result of poor correlation of mobility reduction and contact reduction.

In conclusion, our study illustrates how the disruption of the seasonal pattern of endemic infectious diseases following the COVID-19 NPIs can be used to quantify the factors that govern seasonality. Immunisation strategies for RSV are unlikely to substantially alter the timing of RSV season in Australia dramatically, but the type of vaccination strategy can influence the amplitude of the seasonal incidence. However, these results may not hold for RSV transmission in climate zones with a weaker seasonality, where the shifts in incidence might possibly be more pronounced following the introduction of a vaccine.

## Data Availability

The R/Julia code as well as all input and output data are available in a Github repository (https://github.com/fkrauer/RSV-seasonality-public). The original RSV case notification data are available on the MSW MoH website [27,28]. The Google mobility data were downloaded from [30].

## Author contributions

**FK:** conceptualization, methodology, software, validation, formal analysis, data curation, visualization, writing – original draft, writing – review & editing. **TEF**: methodology, software, validation, writing – review & editing. **MK**: methodology, writing – review & editing. **DH**: methodology, writing – review & editing. **MTS**: project administration, funding acquisition, writing review & editing. **CH**: data curation, writing – review & editing. **MJ**: conceptualization, funding acquisition, writing – review & editing. **OW:** project administration, funding acquisition, writing – review & editing. **TH:** project administration, funding acquisition, writing – review & editing. **SF**: project administration, conceptualization, methodology, funding acquisition, supervision, writing review & editing.

## Ethics

No ethical approval was needed as all data used were anonymised and publically available.

## Acknowledgments

We thank Dr. Christopher Rackauckas, Dr. Seth Axen and the Julia language community on Slack for technical advice.

