## supplemental material for "Estimating RSV seasonality from pandemic disruptions: a modelling study"

### 1. Methods

#### Data

The weekly incidence data are available in PDF files from the NSW Ministry of Health website [1,2]. RSV data until March 2020 are published in the weekly/monthly influenza surveillance reports, data from May 2020 onwards are published in the weekly COVID-19 surveillance reports. No RSV data were reported between 5. April and 3. May 2020 and between 18. July 2021 and 25. July 2021. The data were automatically extracted from the PDF files using tabula (https://tabula.technology/) and manually corrected in case of misalignment or failed extractions. The data on bronchiolitis cases were extracted from the corresponding graphs with Webplotdigitizer (see https://apps.automeris.io/wpd/). The Kenyan attack rate data were extracted from the published paper [3]. The numerators are from table 1 in the main paper, the denominators are given in table S1 in the supplement.

#### Model structure

| Level | State | | Transmission and transitions |
|---|---|---|---|
| 0 | $\dot{S_0}$ | = | $\mu N - \lambda_0(t)S_0 - \mu S_0$ |
| | $\dot{E_0}$ | = | $\lambda_0(t)S_0 - \sigma E_0 - \mu E_0$ |
| | $\dot{I_0}$ | = | $\sigma E_0 - \gamma_0 I_0 - \mu I_0$ |
| | $\dot{R_0^1}$ | = | $\gamma_0 I_0 - 2 * \omega R_0^1 - \mu R_0^1$ |
| | $\dot{R_0^2}$ | = | $2 * \omega R_0^1 - 2 * \omega R_0^2 - \mu R_0^2$ |
| 1 | $S_1$ | = | $2 * \omega R_0^2 - \lambda_1(t)S_1 - \mu S_1$ |
| | $\dot{E_1}$ | = | $\lambda_1(t)S_1 - \sigma E_1 - \mu E_1$ |
| | $\dot{I_1}$ | = | $\sigma E_1 - \gamma_1 I_1 - \mu I_1$ |
| | $\dot{R_1^1}$ | | $\gamma_1 I_1 - 2 * \omega R_1^1 - \mu R_1^1$ |
| | $\dot{R_1^2}$ | = | $2 * \omega R_1^1 - 2 * \omega R_1^2 - \mu R_1^2$ |
| 2 | $\dot{S_2}$ | = | $2 * \omega R_1^2 - \lambda_2(t)S_2 - \mu S_2$ |
| | $\dot{E_2}$ | = | $\lambda_2(t)S_2 - \sigma E_2 - \mu E_2$ |
| | $\dot{I_2}$ | = | $\sigma E_2 - \gamma_2 I_2 - \mu I_2$ |
| | $\dot{R_2^1}$ | = | $\gamma_2 I_2 - 2 * \omega R_2^1 - \mu R_2^1$ |
| | $\dot{R_2^2}$ | = | $2 * \omega R_2^1 - 2 * \omega R_2^2 - \mu R_2^2$ |
| 3 | $\dot{S_3}$ | = | $2 * \omega (R_2^2 + R_3^2) - \lambda_3(t)S_3 - \mu S_3$ |
| | $\dot{E_3}$ | = | $\lambda_3(t)S_3 - \sigma E_3 - \mu E_3$ |
| | $\dot{I_3}$ | = | $\sigma E_3 - \gamma_3 I_3 - \mu I_3$ |
| | $\dot{R_3^1}$ | | $\gamma_3 I_3 - 2 * \omega R_3^1 - \mu R_3^1$ |
| | $\dot{R_3^2}$ | = | $2 * \omega R_3^1 - 2 * \omega R_3^2 - \mu R_3^2$ |
| Cumulative incidence | $\dot{C_0}$ | = | $\sigma E_0$ |
| | $\dot{C_1}$ | = | $\sigma E_1$ |
| | $\dot{C_2}$ | = | $\sigma E_2$ |
| | $\dot{C_3}$ | = | $\sigma E_3$ |

(1)

#### Seasonal transmission

The seasonal forcing of the transmission rate was calculated with a modified Von Mises function of the form

$$\beta_{eff} = \beta_0 + \eta \left( \frac{\exp\left(\frac{cos\left(\frac{2\pi(t-\varphi)}{365}\right)-1}{k^2}\right) - \exp\left(\frac{cos(\pi)-1}{k^2}\right)}{1-\exp\left(\frac{cos(\pi)-1}{k^2}\right)} \right) \tag{2}$$

Where $\beta_0$ is the baseline (minimum) transmission rate, $\eta$ is the amplitude of the peak and $k$ scales the width of the peak. The circular Von Mises distribution is a close approximation to the wrapped normal. Depending on the value of $k$, it can take any shape between a uniform distribution and a sharp, narrow peak. Practically, we limited the upper bound of the prior to two, which allows for any shape between a cosine-like and a sharp, narrow peak. In the case of $\eta = 0$, the seasonal forcing disappears and the transmission rate becomes constant.

The present parameterisation is based on a modified version of the von Mises function (MVM) [4] where

$$MVM = \exp\left(\frac{cos\left(\frac{2\pi(t-\varphi)}{365}\right)-1}{k^2}\right) \tag{3}$$

The maximum of the MVM is 1, the minimum depends on the value of $k$. To have a better control over the minimum and maximum of the seasonal forcing, we re-scaled the MVM according to the following general function:

$$x_{scaled} = target_{min} + (target_{max} - target_{min})\,\frac{x - x_{min}}{x_{max} - x_{min}} \tag{4}$$

Where $target_{min}$ and $target_{max}$ are the desired minimum and maximum of the function x. We define $\beta_0$= $target_{min}$ and $\eta$=amplitude, i.e. $target_{max}$ - $target_{min}$. The MVM is minimal when $t$ corresponds to $\varphi$ minus half a period:

$$x_{min} = \exp\left(\frac{cos\left(\frac{2\pi\left(\varphi + \frac{365}{2} - \varphi\right)}{365}\right)-1}{k^2}\right) \tag{5}$$

which simplifies to

$$x_{min} = \exp\left(\frac{cos(\pi)-1}{k^2}\right) \tag{6}$$

Finally, substituting equation (6) into equation (4) yields equation (2).

#### Force of infection

The force of infection (FOI) among the naïve (i.e. never infected) was calculated as

$$\lambda_0 = \beta_{eff}\left(\frac{I_0 + I_1 + I_2 + I_3}{N}\right)i \qquad (7)$$

where *i* is the reduction of the FOI during the COVID-19 pandemic due to contact and mobility restrictions (see below).

Among the susceptibles of the levels 1-3, the corresponding FOI was scaled relative to the FOI of the preceding level with

$$\begin{aligned} \lambda_1 &= \lambda_0\delta_1 \\ \lambda_2 &= \lambda_0\delta_1\delta_2 \\ \lambda_3 &= \lambda_0\delta_1\delta_2\delta_3 \end{aligned} \qquad (8)$$

#### Observation models

We calibrated the model jointly to two datasets: 1) a time series dataset of weekly incident, laboratory confirmed RSV cases from NSW, and 2) yearly attack rates for symptomatic and asymptomatic RSV from a longitudinal household study from Kenya.

The simulated weekly incident cases for each level *i*, $inc_i$, in the model were calculated as the differences of the cumulative cases for each level of reinfection. Using the level-specific incidences we multiplied the symptomatic incidence in each level by the proportion considered symptomatic, (1-$p_i$):

$$inc\_symp_i = inc_i * (1 - p_i) \qquad (9)$$

The proportion asymptomatic ($p_i$) were taken from a household study in Kenya, where all household members were tested for RSV twice weekly regardless of symptoms [3]. To convert the age-specific measures in the original study to level-specific measures, we assumed that individuals in level 0 corresponded approximately to age 0-1 year olds, individuals in level 1 and 2 corresponded to ages 1-4 and individuals in level 3 corresponded to 5+ year olds.

The total reported cases were then calculated as:

$$inc\_rep = \sum_{i=0}^{2} inc\_symp_i \qquad (10)$$

We assumed the observed reported cases, *inc_obs*, at each timepoint *t* followed a negative binomial distribution

$$inc\_obs_t \sim Negative\ Binomial(r, p) \qquad (11)$$

to account for overdispersion in the observations. We followed the approach of Lindén and Mäntyniemi [5] to reparametrize the negative binomial distribution in terms of the mean-variance relationship instead of the *r*th success with success probability *p*.

Assuming a quadratic mean-variance relationship, the equation for the likelihood becomes

$$inc\_obs_t \sim Negative\ Binomial\left(\frac{1}{\psi}, \frac{1}{\psi\mu_t}\right) \quad (12)$$

where $\psi$ =overdispersion parameter and $\mu$=expectation (i.e. the simulated reported incidence). The overdispersion parameter was fitted, and the expectation was calculated as

$$\mu_t = inc\_rep_t * \rho_t \quad (13)$$

where $inc\_rep_t$ is the simulated reported incidence and $\rho_t$ is the fitted reporting rate.

The reporting rate $\rho$ was assumed to increase over time following a sigmoid function. The instantaneous reporting rate $\rho_t$ was calculated as

$$\rho_t = \rho_0 * exp\left[ln\frac{1}{\rho_0} * (1 - exp^{-q*t})\right] \quad (14)$$

where the rate of increase, $q$, and the initial reporting rate $\rho_0$ were estimated from the data.

The level-specific average attack rates were calculated for each level of reinfection, *i*, every pre-pandemic year, *j*, as

$$AR_{ij} = \frac{cumul_{ij}}{pop_{ij}} \quad (15)$$

Where $cumul_{ij}$ are the cumulative cases in level *i* at the end of year *j* and $pop_{ij}$ is the total population in each level *i* at the end of year *j*. Since the disease transmission was assumed to be in stable periodicity pre-pandemic, the average yearly attack rates are identical for each pre-pandemic year.

We assumed the observed cumulative cases in the household study were a realization of a Binomial distribution:

$$cumul\_obs \sim Binomial(pop\_obs, AR) \quad (16)$$

With *pop_obs* being the total observed population of the household study and AR the attack rate estimated from the model.

The final, joint likelihood was calculated as:

$$Pr\left( inc\_obs_t, cumul\_obs_j \middle| \vartheta\right) = \prod_{t=1}^{T} \Pr(inc\_obs_t|\vartheta) \prod_{j=1}^{J} \Pr(cumul\_obs_j|\vartheta) \quad (17)$$

where $\Pr(inc\_obs_t|\vartheta)$ and $\Pr(cumul\_obs_j|\vartheta)$ are the negative binomial and the binomial probability mass functions defined in equations (13) and (17).

#### Sensitivity analysis

We performed three sensitivity analyses: 1) a fit to a pre-pandemic subset of the data only, 2) a cosine forcing function instead of a Von Mises function, and 3) a fit to the full time series data but without the attack rate data. The cosine function takes the form:

$$\beta_{eff} = \beta_0 \left(1 + \eta \cos \frac{\pi(t - \varphi)}{365}\right) \tag{18}$$

where $\beta_0$ is the average transmission rate and $\eta$ scales the amplitude.

**Description of RSV data**

**Figure S1.** A) Weekly detected and reported RSV cases (black line) and weekly number of samples tested for influenza (blue line) in 2010-2022 in NSW. The red vertical line denotes the implementation of the national lockdown mid-March 2020. B) Weekly counts of visits to the Emergency Departments in NSW for all ages for the years 2010-2019 (data digitized from [2]). C) relative distribution of confirmed RSV cases by month and year. D) Seasonality of RSV cases 2010-2019 ("pre-pandemic") and 2020-2022 ("pandemic").

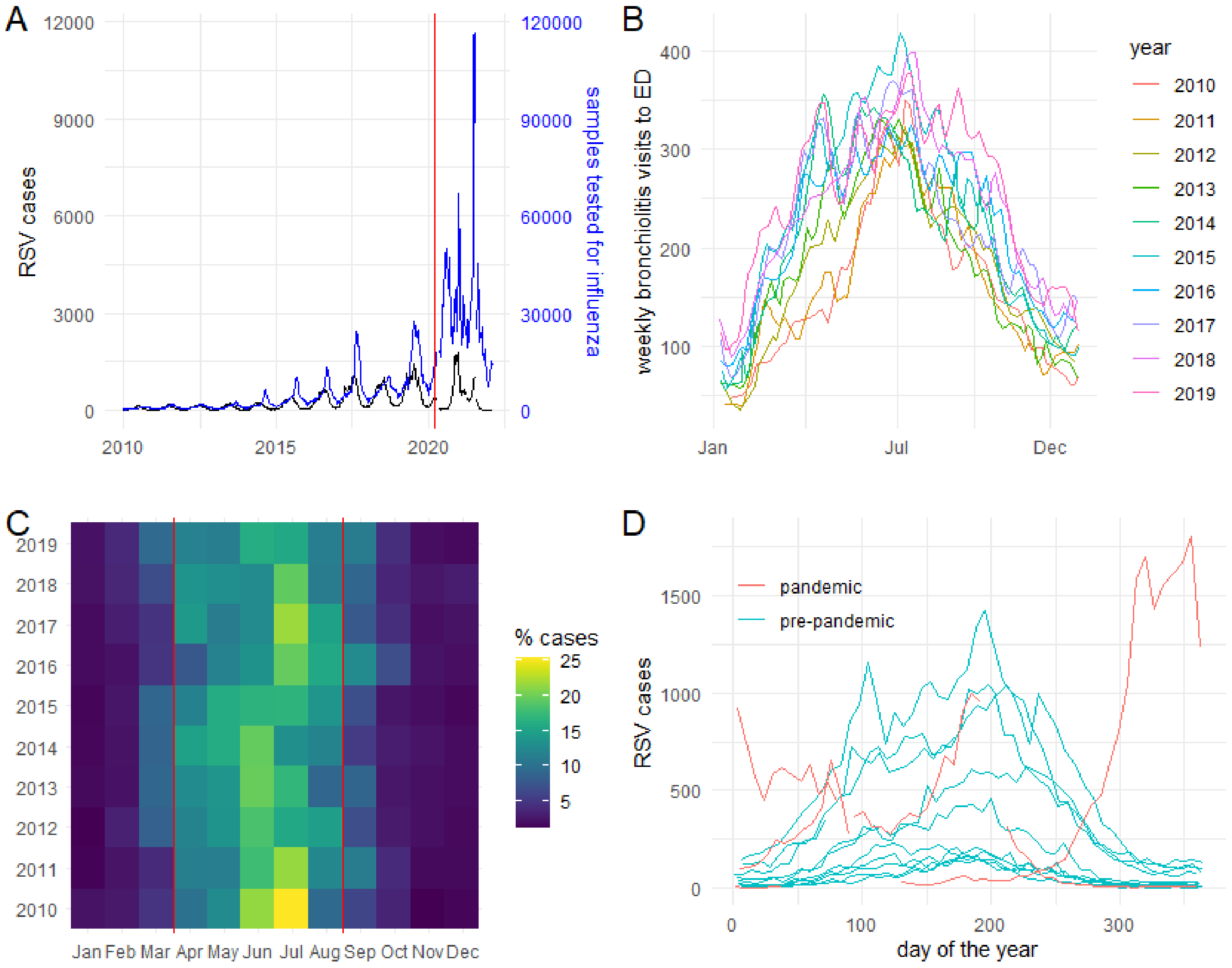

#### Transmission reduction during the pandemic period

For the estimation of the state-level averaged reduction in the FOI we used Google Community Mobility data [6]. These indicators approximate the daily relative reduction in movement measured in different geographical points of interest during the pandemic compared to the baseline of January 3 to February 6, 2020. The data are grouped in six categories (grocery and pharmacy, parks, residential, retail and recreation, transit station and workplaces). We hypothesised that the majority of transmission occurs indoors in places where people spend most time such as workplaces, at home and at schools. The latter is not covered explicitly by Google Community Mobility data. We therefore chose the mobility reduction at workplaces to scale the FOI. For this, we calculated the rolling 7-day median from all available data points. The resulting, averaged mobility index (Figure S2a) was then rescaled between 0 and 1 with the pre-pandemic baseline in March 2020 fixed at 1. This scaling factor, $i$, was then multiplied with the FOI to estimate the effective transmission during the pandemic period.

We also compared the changing movement patterns observed in the Google Mobility reports to the government imposed restrictions in NSW approximated by the Oxford Stringency index [7]**.** As shown in Figure S2b, the mobility data correlated well with the stringency index for workplaces with a stationary effect on mobility reduction for stronger stringency measures. The mobility data also showed the same relationship with the stringency index at schools (Figure S2c), which suggests that workplace mobility reduction may be an acceptable approximation of contact reduction both in adults and children.

**Figure S2.** A) Google Community Mobility index data for locations designated as workplaces (black dots) and rolling 7-day median (red line). B) Agreement of mobility data and fitted contact reductions in NSW based on multiple surveys. The data for the contact reduction were taken from Figure 1 in [8]. C and D) Comparison of scaled mobility data and Oxford stringency index [7] for workplaces and schools.

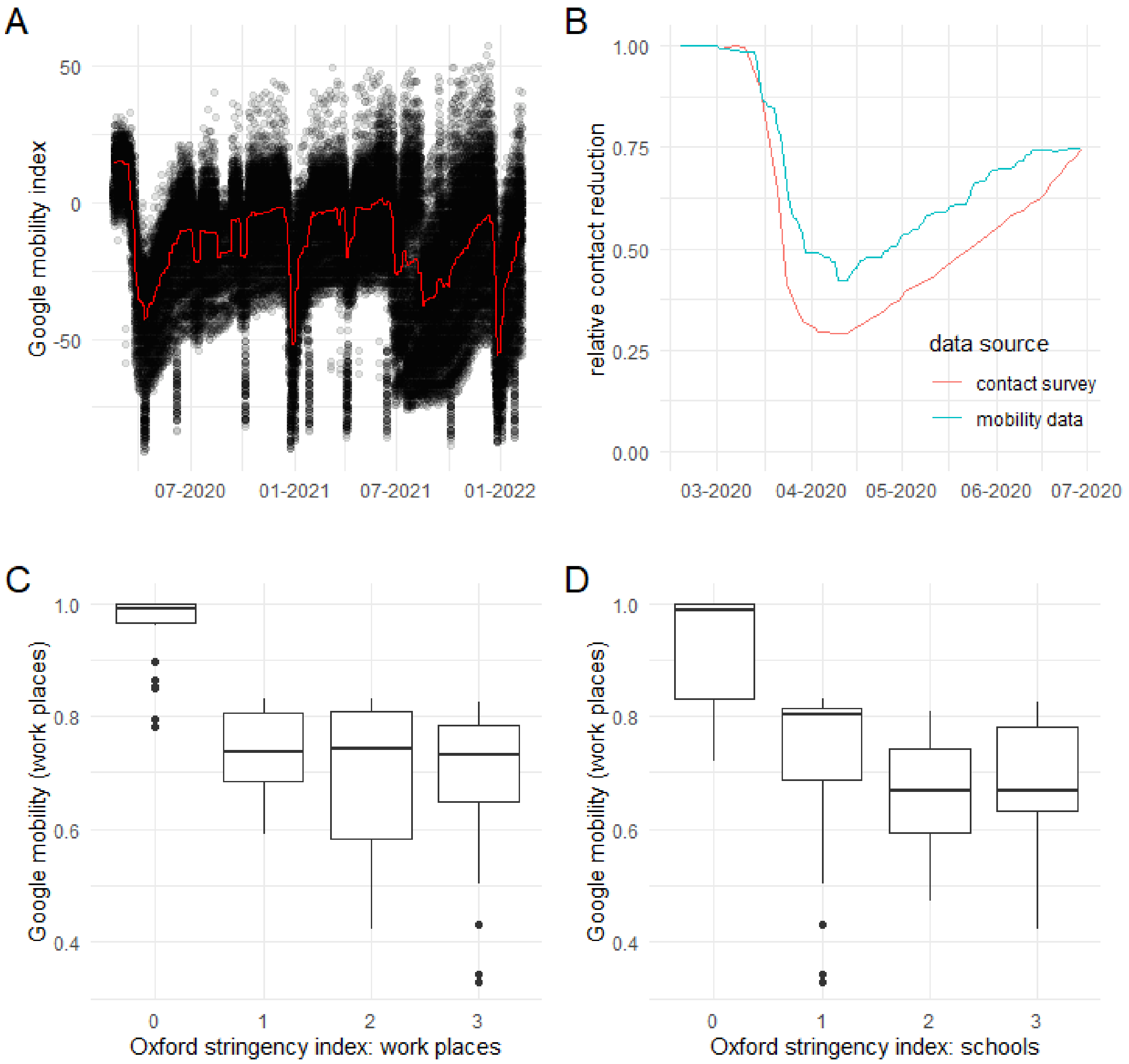

#### Vaccination

Both continuous and pulse vaccination were simulated only for the previously unexposed (level 0). For the continuous strategy, we assumed that new-borns were vaccinated with coverage $v$ immediately after birth and proceeded directly to the compartment $R_0$. The rest is born into the $S_0$ compartment. For the seasonal vaccination, we assumed vaccination of all new-borns as for continuous vaccination plus vaccination of all individuals on level 0 who have not developed prior immunity yet. These are shifted to the compartment $R_0$ at a vaccination rate $\varepsilon$. The vaccination rate is a function of the coverage ($v$) and the duration of the pulse ($d$) and was calculated as

$$\varepsilon = \frac{\log(1-v)}{d} \tag{19}$$

Continuous and pulse vaccination were modelled as follows:

| Level | State | | Transmission and transitions | Continuous vaccination | Seasonal vaccination |
|---|---|---|---|---|---|
| 0 | $\dot{S_0}$ | = | $\lambda_0(t)S_0 - \mu S_0$ | $+ (1-v)\mu N$ | $+ (1-v)\mu N - \varepsilon S_0$ |
| | $\dot{E_0}$ | = | $\lambda_0(t)S_0 - \sigma E_0 - \mu E_0$ | | $-\varepsilon E_0$ |
| | $\dot{I_0}$ | = | $\sigma E_0 - \gamma_0 I_0 - \mu I_0$ | | $-\varepsilon I_0$ |
| | $\dot{R_0^1}$ | = | $\gamma_0 I_0 - \omega R_0 - \mu R_0^1$ | $+ v\mu N$ | $+ v\mu N + \varepsilon(S_0 + E_0 + I_0)$ |

#### 2. Results

##### MCMC Diagnostics

For the main results, we ran ten independent HMC MCMC chains with different seeds, of which all converged to consistent and unimodal posterior distributions within 24 hours. The Bayesian Fraction of Missing Information (BFMI) was >0.84 for all chains suggesting thorough exploration of the posterior. The trace plots show good mixing of the chains. ESS ranged from 878-2475 across all fitted parameters. Rhat estimates were <1.01 for all parameters suggesting chain convergence.

##### Additional quantitative results

The reduction of susceptibility compared to level 0 was 0.83 (95% Cr 0.73-0.0.90), 0.77 (95% CrI 0.62-0.89) and 0.24 (95% CrI 0.17-0.32) for levels 1, 2 and 3, respectively. These estimates are in line with results from another modelling study [9]. Assuming a constant mortality rate, a rectangular population structure and a life expectancy of 80 years, we also calculated the proportional distribution of cases among the infectious compartments and the corresponding average age at infection. The first infection is experienced at age 0.5 (95% PPI 0.3-0.6) years, the second infection at age 2.1 (95% PPI 1.9-2.3) years and the third infection at age 3.8 (95% PPI 3.6-4.0) years.

**Figure S3**. Trace plots and density plots of the posterior samples. A) Fitted to the full dataset (2010-2022) with the Von Mises forcing. B) fitted to the pre-pandemic data (2010-2019) with the Von Mises forcing.

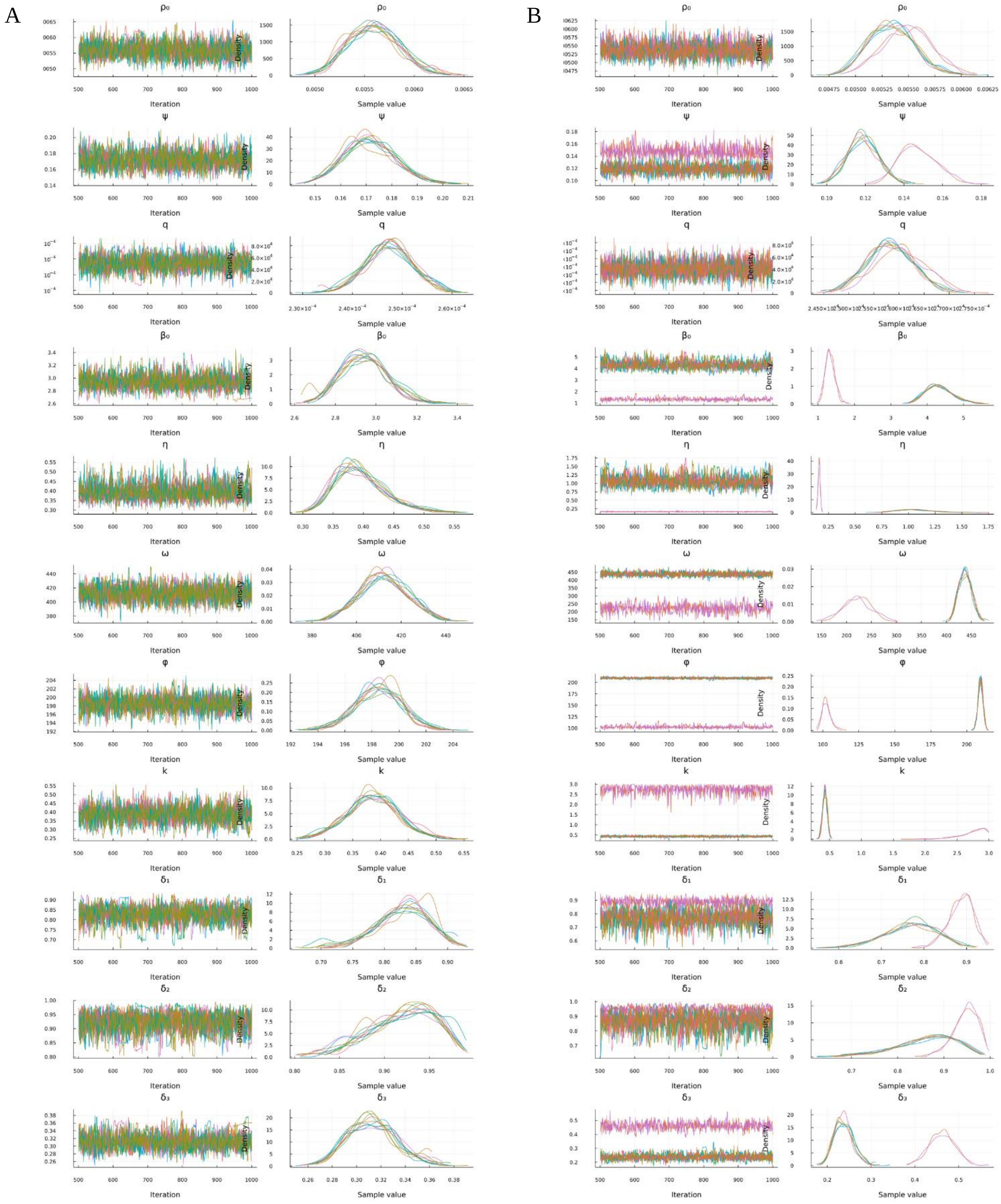

**Figure S3 continued**. Trace plots and density plots of the marginal posteriors C) Fitted to the full time series dataset (2010-2022) with the cosine forcing. D) fitted to the full time series dataset (2010-2022) with the Von Mises forcing but excluding the attack rate data from Kenya.

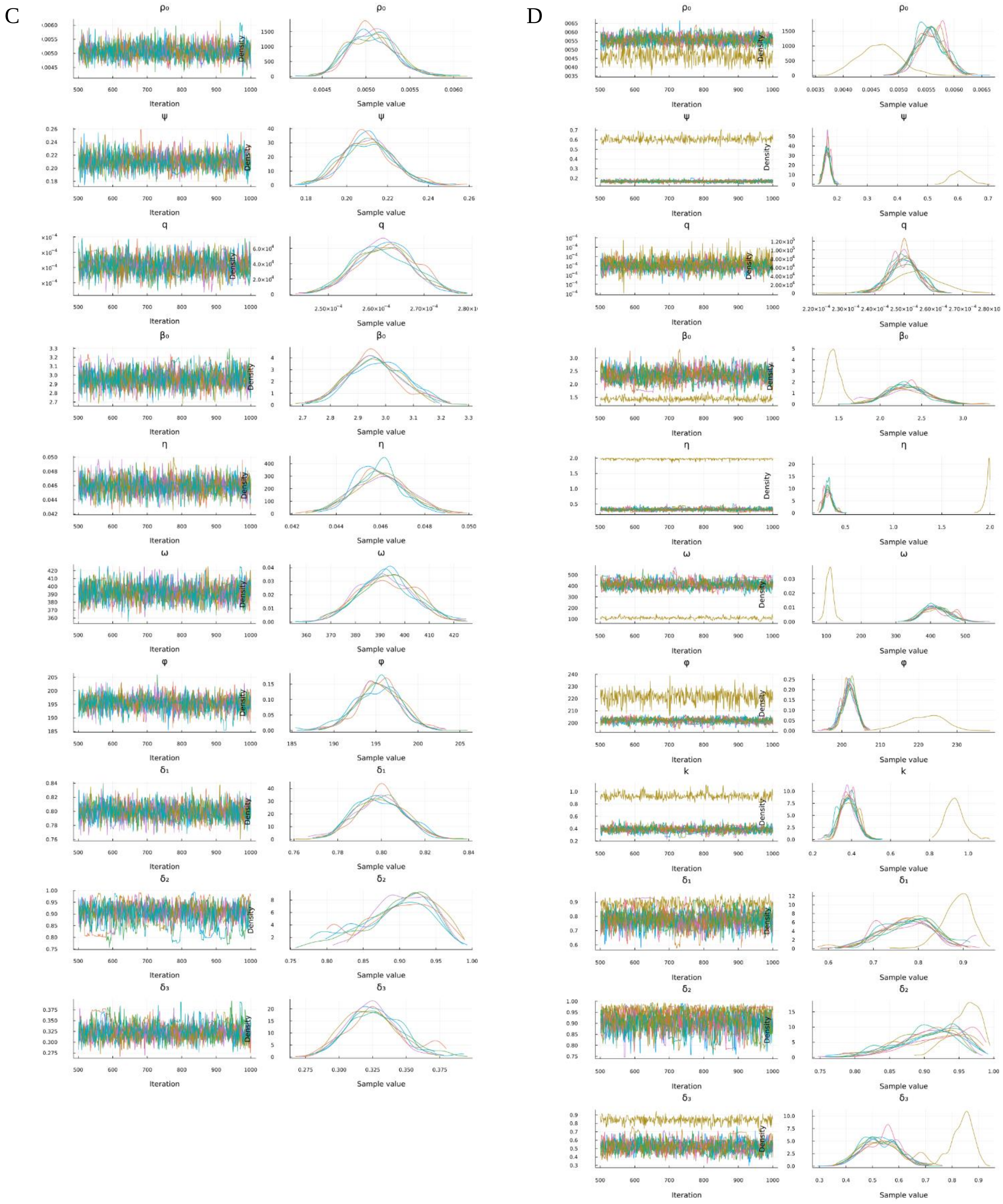

**Figure S4.** Log posterior plots for the four fitted models. A) Mises 2010-2022 with joint likelihood, B) Mises 2010-2019 with joint likelihood, C) Cosine model 2010-2022 with joint likelihood, D) Mises 2010-2022 without AR data.

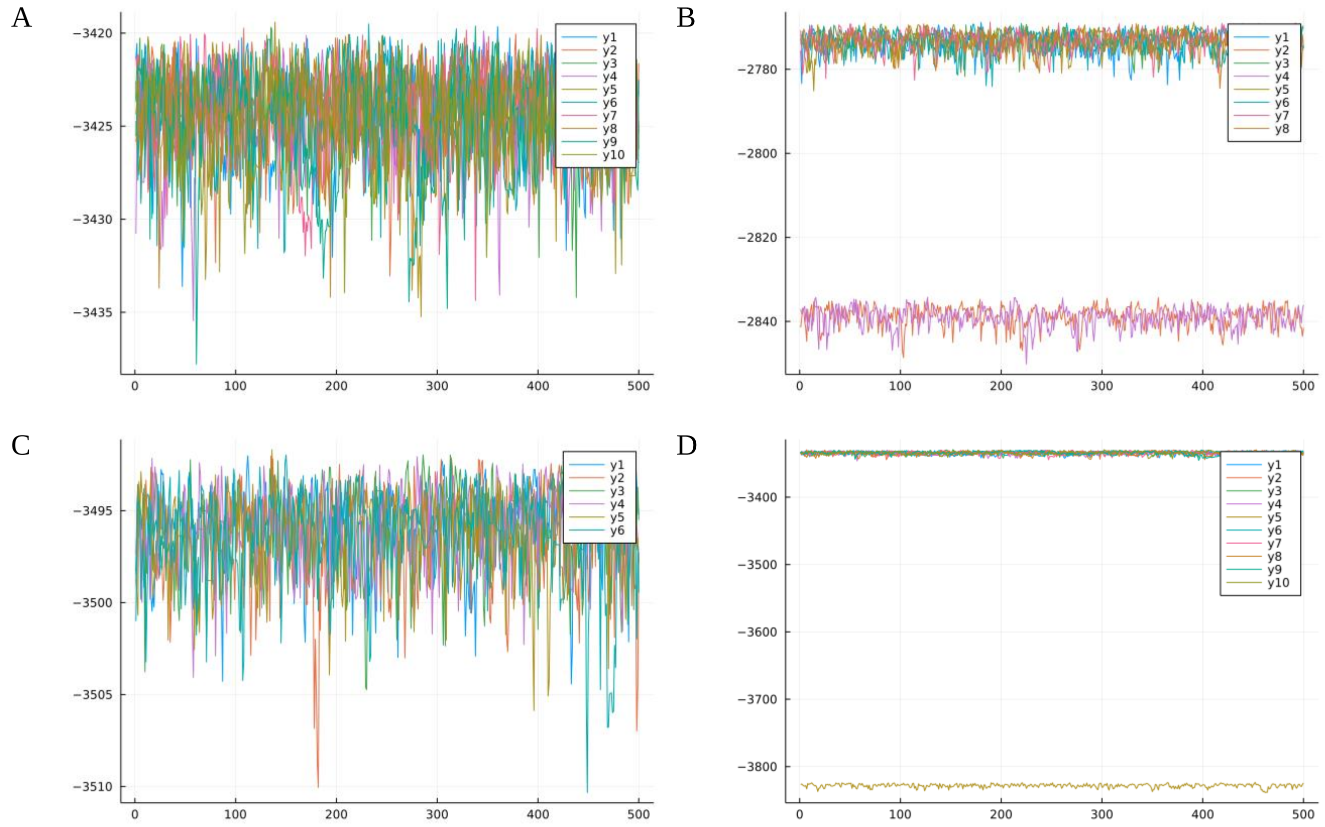

**Figure S5.** Comparison of fit to the full time series (2010-2022) and fit to the pre-pandemic data (2010-2019) only. The two chains stuck in a local mode were discarded for this analysis. A) Trajectory fit of the model fitted to the full dataset (black lines) and the pre-pandemic dataset (red line and ribbon). The black dots are the data points. B) Cross-correlation of posterior parameters using Pearson correlation coefficient's. Red borders indicate a correlation > 0.7 or < -0.7. C) Posterior densities of the pre-pandemic (red) and the full dataset (blue).

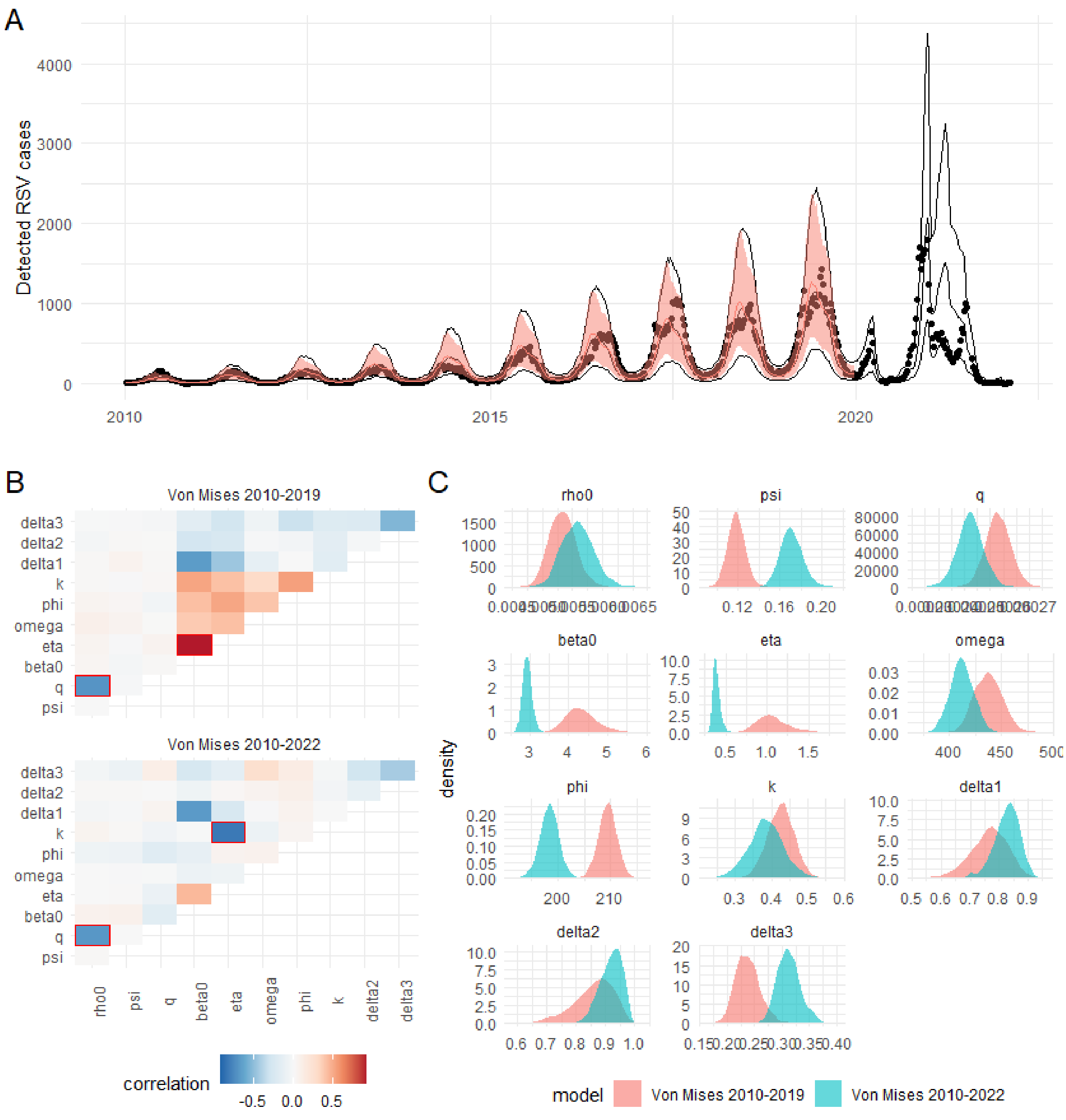

**Figure S6**. Results of the sensitivity analysis using a cosine forcing instead of a modified Von Mises function. A) The trajectory fit based on the cosine forcing (red shaded area) is similar to the main modelling results (black lines) but has a larger uncertainty. B) Both the baseline (off-season) transmission rate as well as the seasonal peak are marginally lower for the cosine forcing (red line and ribbon) than the Von Mises forcing (black lines). C) The marginal posterior distributions from the cosine forcing function (red) are roughly consistent with the main results (blue). The duration of immunity was estimated slightly shorter and the dispersion parameter of the observation model is slightly larger for the cosine model. Note that the parameters $\eta$ and $\beta_0$ were omitted due to the different interpretation in both models, and k was omitted as there is no equivalent in the cosine model.

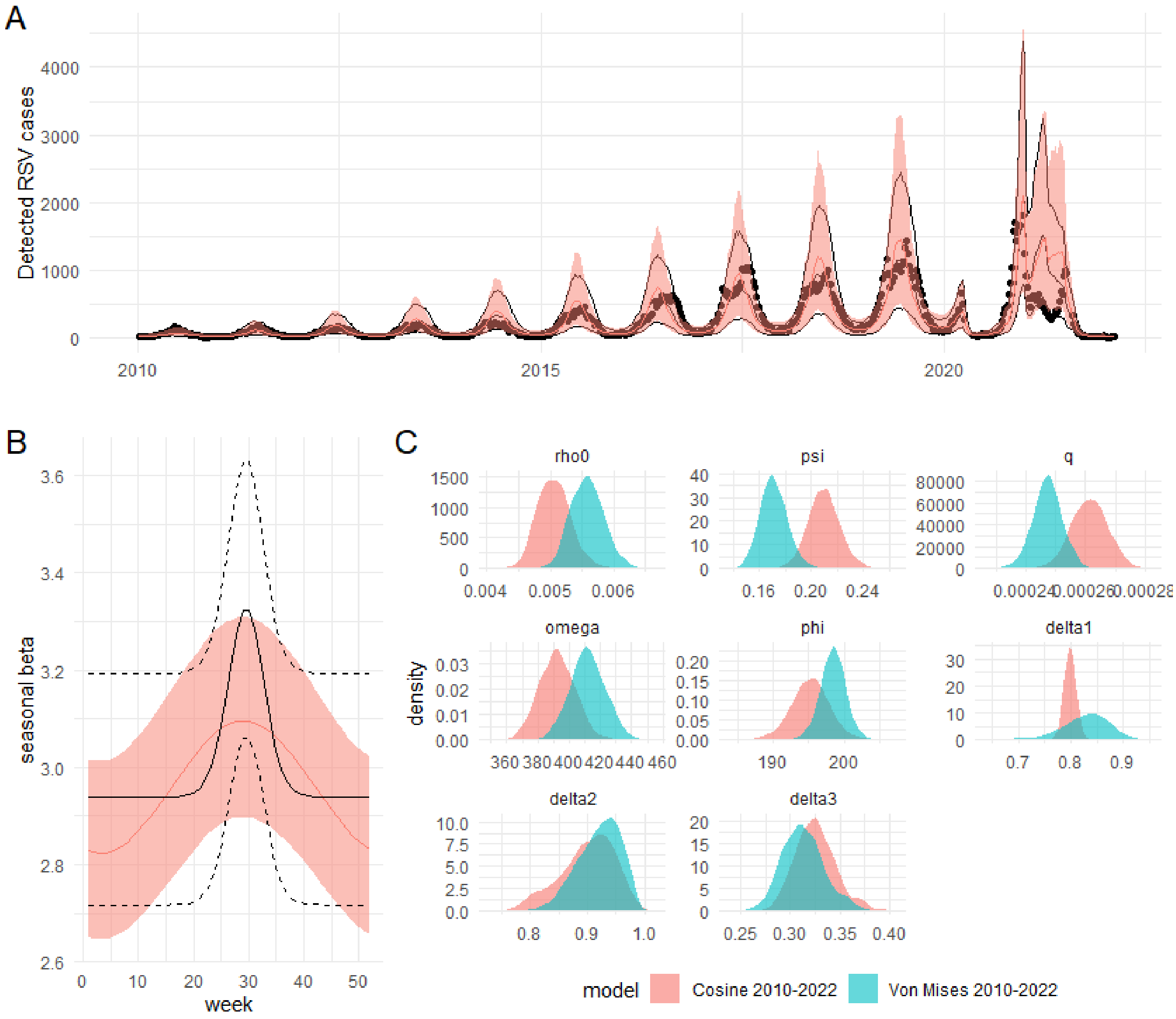

**Figure S7**. Results of the sensitivity analysis: Von Mises model fitted only to the weekly time series data from NSW ignoring the attack rate data from Kenya. The chain stuck in a local mode was discarded for this analysis. A) The trajectory fit of the model fitted to the joint dataset (black lines) was identical to the fit to the time series data only (red line and ribbon). The black dots are the data points. B) The attack rates for level 3 were closer to the estimates suggested by household study data when the model was fitted to both the time series data and the attack rate data (red boxplots) than when fitted to the time series only (blue boxplots). C) There was an overall stronger cross-correlation of the posterior parameters when the model was fitted only to the time series data (bottom plot), which was partially reduced when the model was fitted jointly (top plot). Red borders indicate a correlation > 0.7 or < -0.7. D) The posterior densities of the fit to the joint datasets (red) were narrower and more informative than from the fit to time series only (blue).

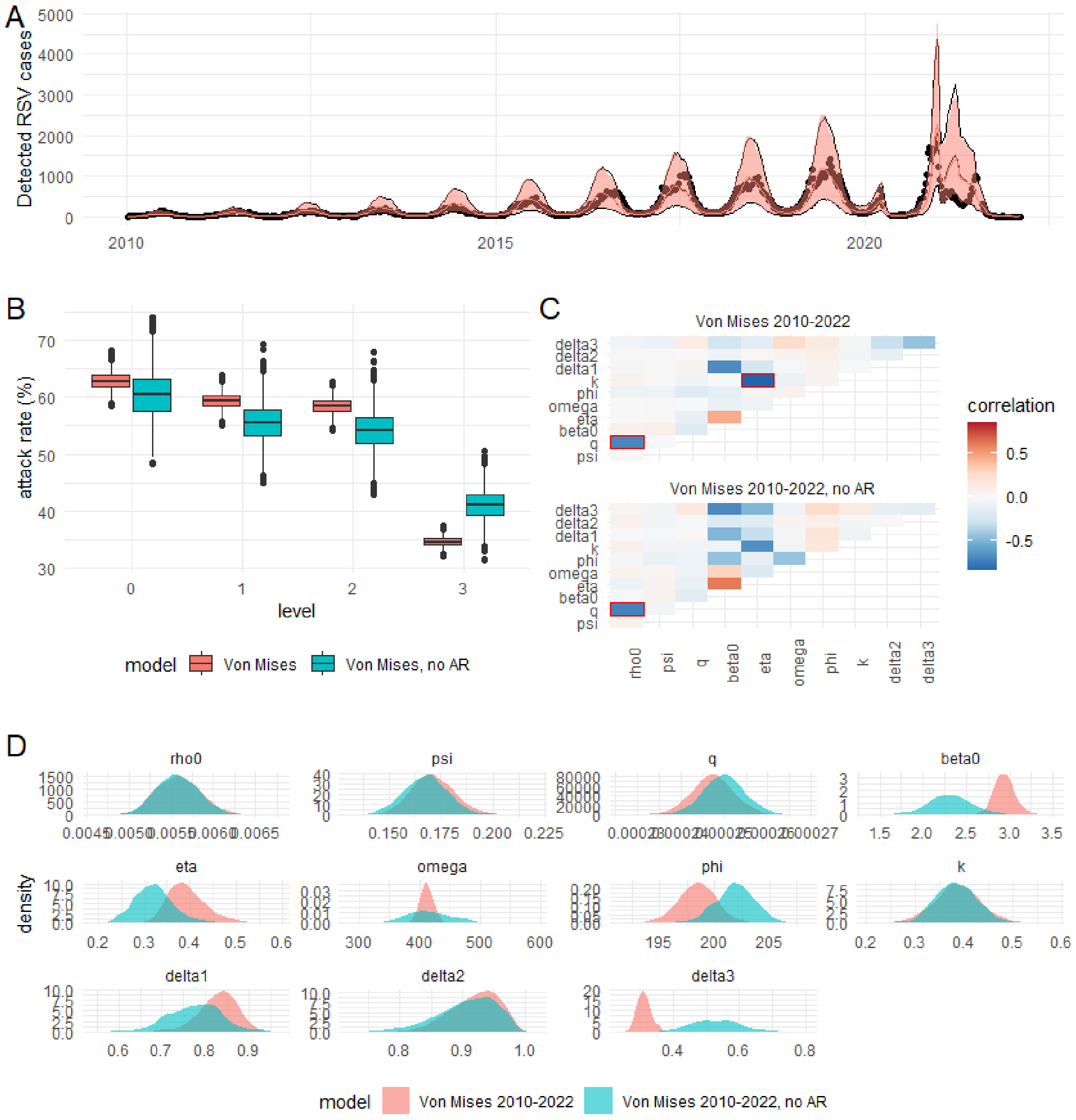
